# How much leeway is there to relax COVID-19 control measures?

**DOI:** 10.1101/2020.06.12.20129833

**Authors:** Sean C. Anderson, Nicola Mulberry, Andrew M. Edwards, Jessica E. Stockdale, Sarafa A. Iyaniwura, Rebeca C. Falcao, Michael C. Otterstatter, Naveed Z. Janjua, Daniel Coombs, Caroline Colijn

**Affiliations:** Pacific Biological Station, Fisheries and Oceans Canada, Nanaimo, BC, Canada; Department of Mathematics, Simon Fraser University, Burnaby, BC, Canada; Department of Biology, University of Victoria, Victoria, BC, Canada; Department of Mathematics and Institute of Applied Mathematics, University of British Columbia, Vancouver, BC, Canada; British Columbia Centre for Disease Control, Vancouver, BC, Canada; School of Population and Public Health, University of British Columbia, Vancouver, BC, Canada

## Abstract

Following successful widespread non-pharmaceutical interventions aiming to control COVID-19, many jurisdictions are moving towards reopening economies and borders. Given that little immunity has developed in most populations, re-establishing higher contact rates within and between populations carries substantial risks. Using a Bayesian epidemiological model, we estimate the leeway to reopen in a range of national and regional jurisdictions that have experienced different COVID-19 epidemics. We estimate the risks associated with different levels of reopening and the likely burden of new cases due to introductions from other jurisdictions. We find widely varying leeway to reopen, high risks of exceeding past peak sizes, and high possible burdens per introduced case per week, up to hundreds in some jurisdictions. We recommend a cautious approach to reopening economies and borders, coupled with strong monitoring for changes in transmission.

---

The novel severe acute respiratory syndrome–coronavirus 2 (SARS-CoV-2 virus), which emerged at the end of 2019, has to date caused a global pandemic with over 7 million confirmed cases of coronavirus disease 2019 (COVID-19) and 408,000 deaths worldwide as of June 9, 2020 [1]. To date, there is no vaccine or cure, and it appears that asymptomatic individuals can be infectious. Accordingly, wide-ranging non-pharmaceutical interventions (NPIs) such as hand hygiene, face masks, physical (social) distancing, banning mass gatherings, and strict lockdowns have been among the primary tools for reducing COVID-19’s spread [2–6].

As a result, incidence in many jurisdictions outside China followed a similar pattern (e.g., Fig. 1B–M). After an initial phase of occasional detection (typically during late January to February and commonly due to imported cases), case counts grew rapidly (typically during early March). At this point, NPIs were put in place, in the form of “lockdowns” or other requirements for social and physical distancing. Case counts generally continued to rise for several weeks until the impact of NPIs became observable as a flattening and then decline of the epidemic curve. The economic, social, and health costs of NPIs have been significant.

**Figure 1:**
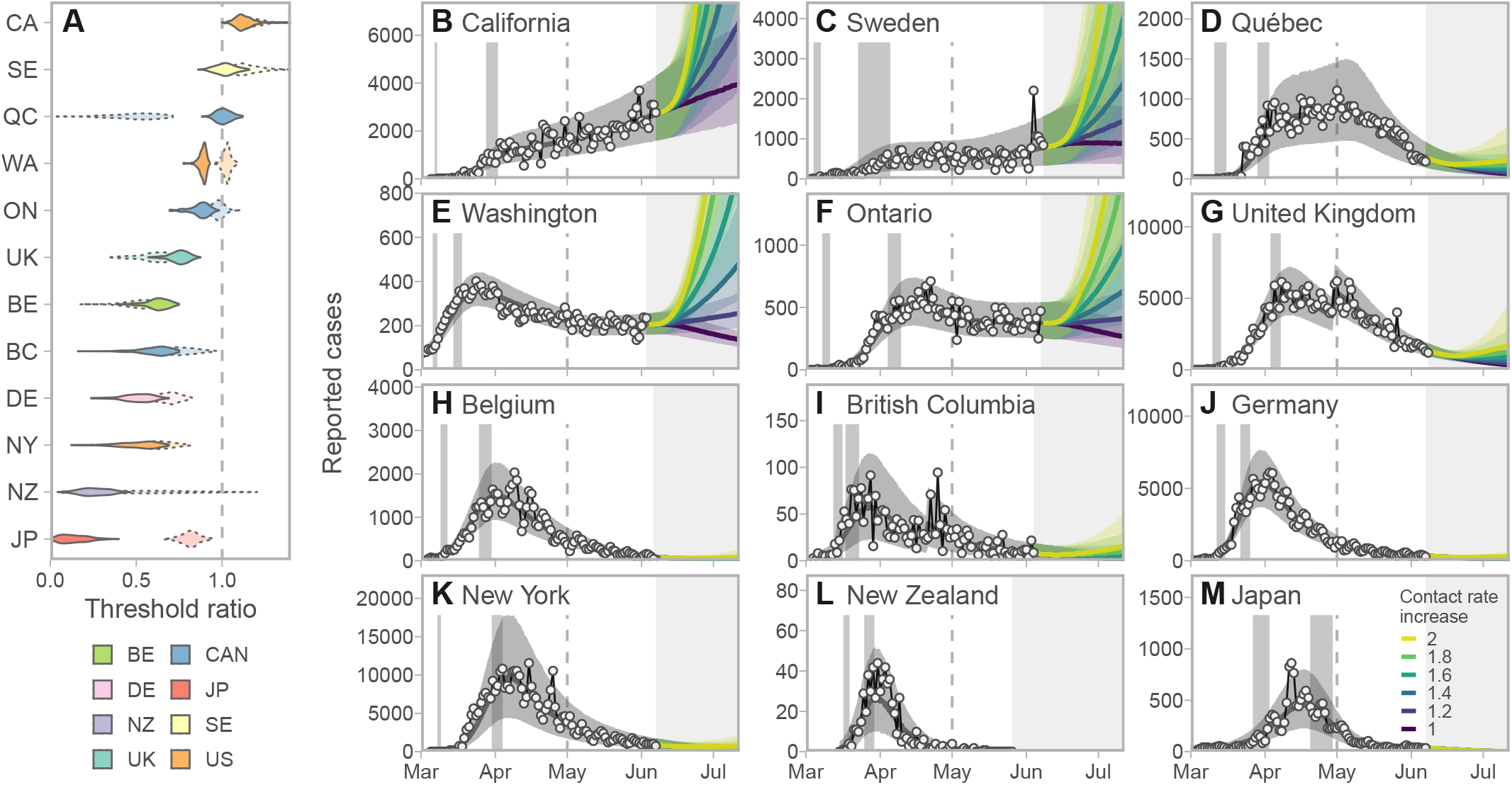
Projected cases given scenarios of relaxed control measures strongly depend on the leeway between the estimated contact rate and the threshold for increase. **A:** Posterior densities of the ratio between the contact rate and the threshold (the value above which exponential increases are expected). Darker violins represent the post-measures period and paler dotted violins represent the recent (post May 1) estimates. Jurisdictions with contacts well below the threshold have more leeway to relax control measures. **B–M:** Model fits and projections at 6 multiplicative contact rate increases, from a baseline from the *lower* of the estimates from the two time periods. Solid lines represent posterior medians and ribbons represent 90% credible intervals. Dots and thin lines represent reported case data. Vertical grey bands indicate 90% credible intervals for the start and end times of initial control measures ramp. Dashed vertical lines indicate the start of the “recent” period (May 1). The choice to project from a baseline of the lower of the post-measures and recent estimates means that projections are based on measures at the stricter time period in all jurisdictions. Regions are arranged by decreasing mean threshold ratio in the immediate post-measures period. per 20,000 given these increases; New York’s previous peak was high and the risk of exceeding it is correspondingly low.

Following declines in incidence, many jurisdictions are now beginning to partially lift restrictions, reopen their economies, and are allowing travel across regional and international boundaries [7–9]. Here, care must be taken not to undo the benefits of widespread NPIs. This is especially true given that large studies undertaken in high-prevalence settings do not indicate that herd immunity has been reached [10]. However, the degree of flexibility, or “leeway”, that exists to increase activity without causing a major resurgence or “second wave” of cases is largely unknown. The flexibility that exists in a given location is dependent on the local circumstances governing transmission, as well as the restrictions that are currently in place [11, 12]. It is essential to estimate the risk associated with increased social and economic activity, and to understand this risk within and between particular jurisdictions, before making decisions around reopening.

We propose that discussions of COVID-19 risk in the context of reopening local economic activity, and of reopening borders and trade, should consider three aspects of transmission dynamics: (1) the probability that infections are rising at the current time in a jurisdiction, even if reported cases are declining; (2) the probability that a given increase in social and economic activity in the general population will lead to a substantial growth in cases over the coming weeks, and (3)—with regards to travel and border reopening—the number of introduced cases and their likely impact in the destination. Using a mathematical model fit to local case data for a selection of jurisdictions with differing epidemics, we estimate the leeway for reopening without causing increasing COVID-19 cases, and the probabilities that reopening will lead to cases increasing above thresholds after a fixed time. The model reflects a portion of the population engaging in distancing and related measures: these individuals are at reduced risk of encountering infectious individuals, and are less likely to be encountered themselves—for example because they are able to work from home, consistently wear masks, or avoid social situations (see Methods). For each of 12 jurisdictions worldwide, selected for their diversity of epidemic trajectories and NPIs, we first estimate the impact to date of widespread NPIs and then calculate how close the estimated contact rate is to the threshold for epidemic growth (Figs 1, S2, S3). We estimate this both in the period immediately following NPI measures (late March to the end of April) and after May 1, as some jurisdictions have already begun to reopen as of the time of writing. We refer to these time frames as “post-measures” and “recent” and use the idea of leeway to describe the room between their current state and the threshold beyond which cases would begin to grow.

We find that after initial NPI measures took effect, some jurisdictions had substantial leeway to re-open (Japan, New Zealand, Germany, New York, British Columbia, and Belgium), with an above-0.99 probability that contact rates were below 80% of the threshold for epidemic growth. Japan and New Zealand had the most leeway, with contact rates well below half the threshold. In contrast, some had little leeway (the United Kingdom (UK), Washington, and Ontario) and some had none, as cases were still rising (Quebec, Sweden, and California). Estimates for the period after May 1 find that some jurisdictions have little or no leeway for further re-opening (California, Sweden, Washington, Ontario) as they are at or above the critical threshold. Some have used part of their leeway already (Japan, Germany, and New York, and British Columbia; Fig. 1A). Several have more leeway than they did immediately after NPI measures took effect (Belgium, the UK, and Quebec, with Quebec now well below the threshold and the UK now with > 0.99 probability of being < 80% of the threshold). New Zealand has so few cases that estimation with this modelling framework leaves considerable uncertainty.

We forecast the impact of relaxing distancing measures by increasing contact rates, starting from a baseline of the lower of the post-measures and recent estimates (Fig. 1B–M). The UK, Belgium, and Quebec moved to stricter control after May 1. All have some leeway as of the time of writing, though increasing contact beyond 60% above the recent estimate would likely lead to a growing epidemic in the UK and Quebec. Belgium has substantial leeway to re-open. The remaining jurisdictions have used some of their leeway already. Those with little to no leeway to begin with now show rapid increases if contact is increased (California, Sweden, Washington, and Ontario). British Columbia had some leeway to re-open and has done so; a doubling of contact compared to the post-measures baseline would likely lead to rises in case numbers. Germany, New York, New Zealand, and Japan show low risks of rising cases. These results are robust to assumptions about the duration of infection and reasonable priors on the fraction of individuals distancing (Fig. S4).

Rising case numbers may be tolerable, depending on the costs and severity of measures needed to keep cases in check, the capacity of the health care system to cope with increases in COVID-19 cases, and a population’s preference regarding the balance of widespread measures vs. increases in incidence. Policy makers could factor into their decision-making the probability and time frame of new cases that may arise following reopening. Fig. 2 shows our estimates of the probability of exceeding the peak number of cases to date, and the probability of reaching 1 incident reported case per 20,000 individuals under different increases in contact rates (again from a baseline of the time period in which control was stricter). Given similar increases in contact rate, Ontario, Washington, Sweden, and California are most likely to exceed both 1 incident case per 20,000 and their historical peaks in the coming 6 weeks. The UK has a small risk of exceeding its previous peak (probability of 0.06 with a doubling of contact rates from the post-measures period). New York, the UK, and Quebec have some risk of exceeding 1 case

**Figure 2:**
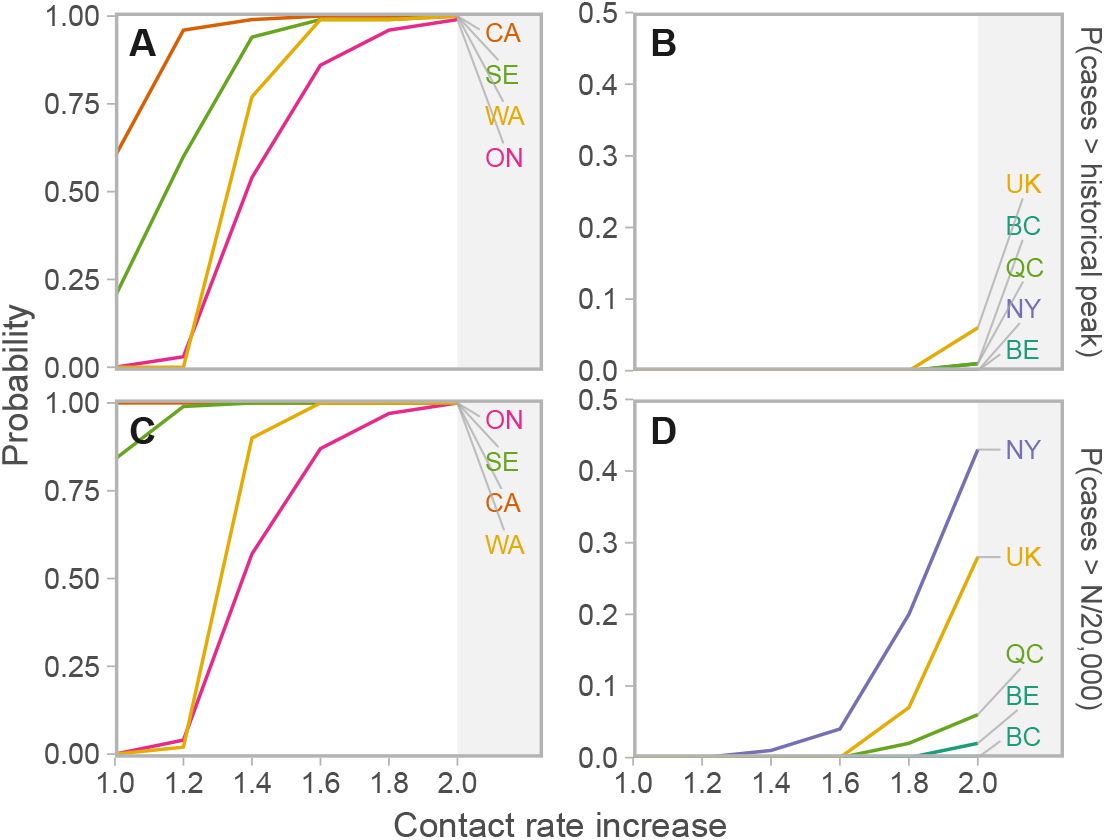
Probabilities of cases exceeding reference thresholds at 6 weeks in the future depend on contact rate increases and jurisdiction. Projections are from a baseline of the lower of the post-measures and recent estimates. **A, B:** Probability of exceeding the historical “first wave” maximum. **C, D:** Probability of reported cases per day exceeding 1/20,000 of the population (*N*). ON: Ontario, WA: Washington, CA: California, QC: Quebec, BC: British Columbia, NY: New York, SE: Sweden, UK: United Kingdom, BE: Belgium, DE: Germany, NZ: New Zealand, JP: Japan.

There is pressure to reopen borders to business and leisure travelers due to the social and economic costs of travel restrictions. We modelled the impact of introducing imported cases at a constant rate to estimate the impact on total cases in each jurisdiction, taking uncertainty in the contact ratios (and other posterior estimated quantities; see Supplementary Information) into account (Fig. 3). Our results illustrate the expected extra cases resulting from one imported case per week over six weeks. Assuming independence of imported cases, these results can be scaled to realistic rates of importation (e.g., for 100 imported cases, multiply expected extra cases by 100). In Japan, where the dynamics are well below the threshold in all posterior samples, each importation results in few additional cases. Meanwhile, in California or Sweden, because there is a high posterior probability that transmission is above the threshold, introduced cases are more likely to cause extended chains of transmission and contribute large case volumes. The result is that up to approximately 100 new cases may result (over six weeks) from a weekly introduction of a single case. Fig. 3 is generated under the assumption that introduced cases join the general population, have access to its testing and control procedures, and engage in its broader distancing and NPI behaviors, making these conservative projections.

**Figure 3:**
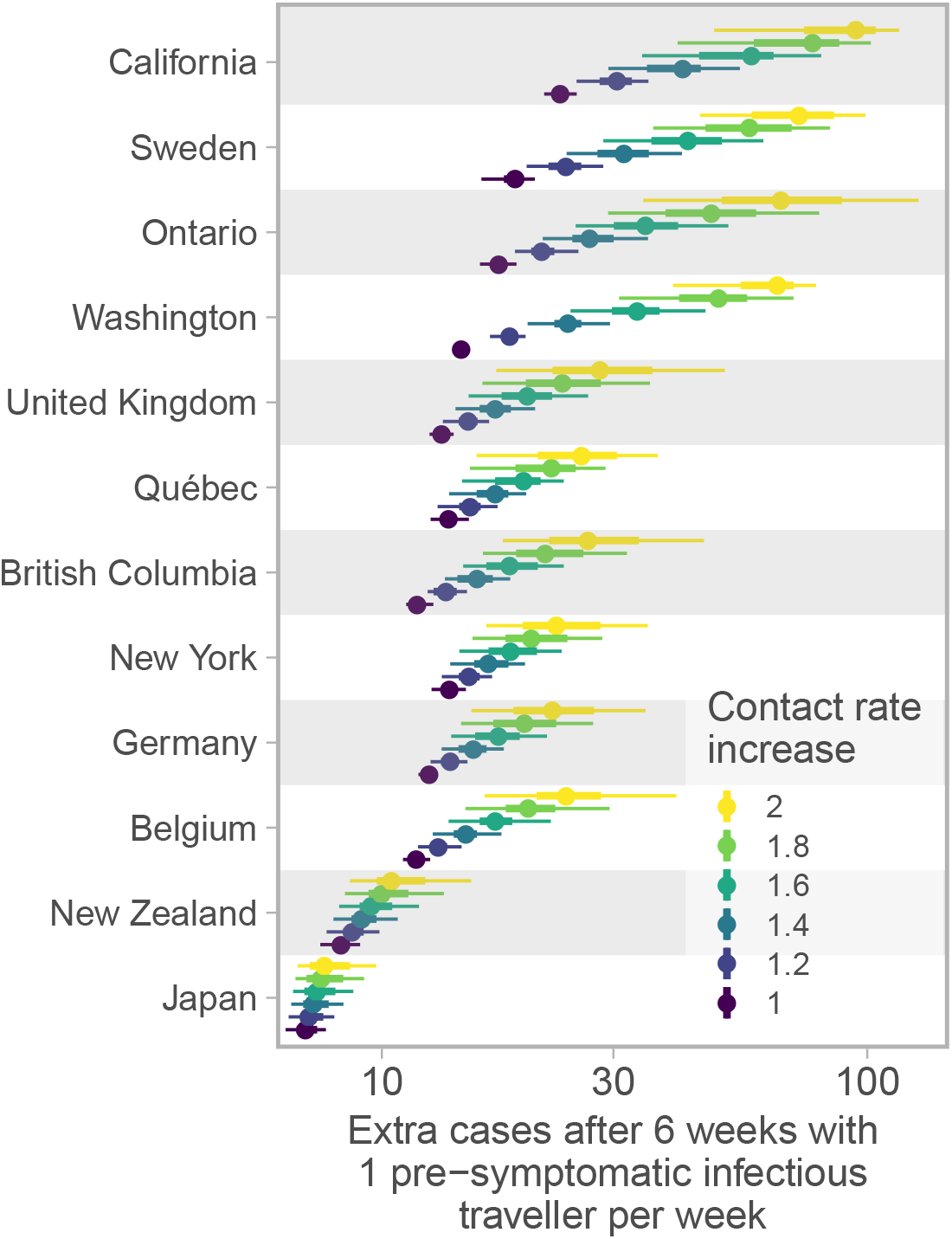
Cases resulting from one import per week over 6 weeks range from fewer than ten to hundreds and depend on contact in the destination population. Dots represent medians and thick and thin line segments represent 50% and 90% credible intervals; the x-axis is log distributed. Contact rate increases are based on the lower of the post-measures and recent contact ratio estimates. Regions are ordered by the average extra cases across contact rate increases. Extra cases are compared to a projection that does not include weekly imports; travelers themselves have not been removed from the totals.

To interpret these results with reference to borders and travel requires consideration of the specific jurisdictions involved. Consider a border opening from jurisdiction A to jurisdiction B. If both jurisdictions are well below their thresholds, then the probability of a large volume of new cases resulting from introductions is low, primarily because general transmission will be prevented in jurisdiction B, but also because prevalence is likely to be low in A, though this depends on the epidemic, testing, reporting, and population dynamics in A. If the destination is near its threshold, then introduced cases could result in exponential growth in B. This effect could be amplified if travelers join a congregate setting or are less socially distanced than the general population due to tourism or work activities, or if they have reduced access to local health care and control measures such as contact tracing. In addition, if jurisdiction A is near its own threshold, then there may be as-yet-unobserved exponential growth of cases in A, affecting the rate of introduction to B. Furthermore, travel itself may result in additional transmissions.

The COVID-19 pandemic has seen an unprecedented number of travel restrictions and border measures, in spite of WHO recommendations against unnecessary closures, weak evidence that these are effective in preventing pandemic influenza [13] (though they do reduce spread and buy time [14]), and concerns about their impact on movement of medical supplies and personnel [15]. There is now some discussion of “travel bubbles” in which countries or jurisdictions experiencing comparable levels of risk open borders to travel and commerce [16]. As jurisdictions with low case numbers move to reopen their economies (likely causing the epidemic threshold as measures are relaxed), they will be at renewed risk of introductions. We suggest that the highest-risk borders arise when a source jurisdiction has prevalent cases and the destination jurisdiction is near or above its threshold, or reopening to the extent that cases could now spread widely despite earlier successes. Due to variations in testing, we cannot know the relative prevalence [17], but we would predict, among the locations in our study, that introductions into California, Sweden, Ontario, and Washington carry the highest risk, followed by the UK. Interactions among these jurisdictions would carry the highest risk, despite that by some indicators the overall COVID-19 control in several of these is similar. Interactions among the UK, Quebec, BC, NY, Germany, and Belgium are lower risk but the probability of causing dozens of new cases per introduced case per week remains considerable. Furthermore, jurisdictions with small historical peaks (e.g., British Columbia, New Zealand) could easily be put in a position of exceeding their historical peak as a result of introduced cases from a region with higher prevalence.

The model and underlying data have limitations. The data are provided by jurisdictions and depend on testing protocols and capacity, delays to reporting, different base populations being tested, and other variations [18]. Indeed, this motivates using inferred summaries like the leeway, in lieu of direct comparisons of case counts. Our approach accounts as much as possible for differences in testing through time, for the local dynamics of distancing behavior, and different starting intensity and timing of different epidemics. However, our model estimates are oriented towards widespread NPI and distancing measures, and implicitly attribute changes in case dynamics to contact rates. In truth, transmission dynamics involve a complex function of outbreak control, management of COVID-19 in health care settings, reduction in community transmission, reporting, contact tracing and other public health measures. Our notion of contact rates combines both rate of interaction and probability of infection during interaction; thus, increased rates of interaction during reopening may, to a certain degree, be possible without increased transmission if key public health measures (e.g., hand hygiene, physical distancing) are strictly adhered to. Our model also assumes a simple population structure—data for more complex populations being largely lacking. In addition, the numbers of reported cases per prevalent case will change as testing is widened, and this is not modelled in our forecasts.

Amidst differing epidemics and control measures, each jurisdiction has a leeway—the room between the current state and the threshold—and this is comparable from place to place. The leeway, together with model fits that are informed by data and which describe the uncertainty in how much leeway there is, can provide a quantitative basis for decisions about reopening. We are at a unique time in this pandemic, with a so-called “first wave” receding not due to immunity, but due to widespread behavioural change. Given that reopening will occur, this leaves populations vulnerable to resurgence of cases, driven both by local transmission and sparked by introductions. Our results indicate that jurisdictions should proceed with great caution, particularly where there is a substantial probability that they are near the threshold already; rapid monitoring should seek signs of increased community transmission and clustered outbreaks as early as possible. To mitigate risks associated with imported cases and reopening borders, it is important to account for the risk of growth in the general population together with the likelihood that imported cases will arrive in high-risk settings. We recommend that policy-makers carefully consider (i) whether imported cases and seeded outbreaks are likely to be identified and managed to the same degree as those in the local population; (ii) whether travellers will engage in high-risk or high-contact activities, especially within marginalized populations; and (iii) whether local trace and test strategies have the capacity to manage imported cases and nascent outbreaks.

## Methods

### Model description

We extend the SARS-CoV-2 susceptible-exposed-infectious-recovered (SEIR) model developed in Ref. [19]. The model allows for self-isolation and quarantine through a quarantine compartment and a reduced duration of infection (compared to the clinical course of disease). We model a fixed portion of the population that is able to participate in physical distancing; each of the SEIR compartments has an analogous compartment in the distancing group (Fig. 4). We extend our model [19] here by estimating additional parameters in a Bayesian context including the timing of the physical distancing ramp, the scale of the initial cases, and multiple contact rates through time for those practicing distancing.

**Figure 4:**
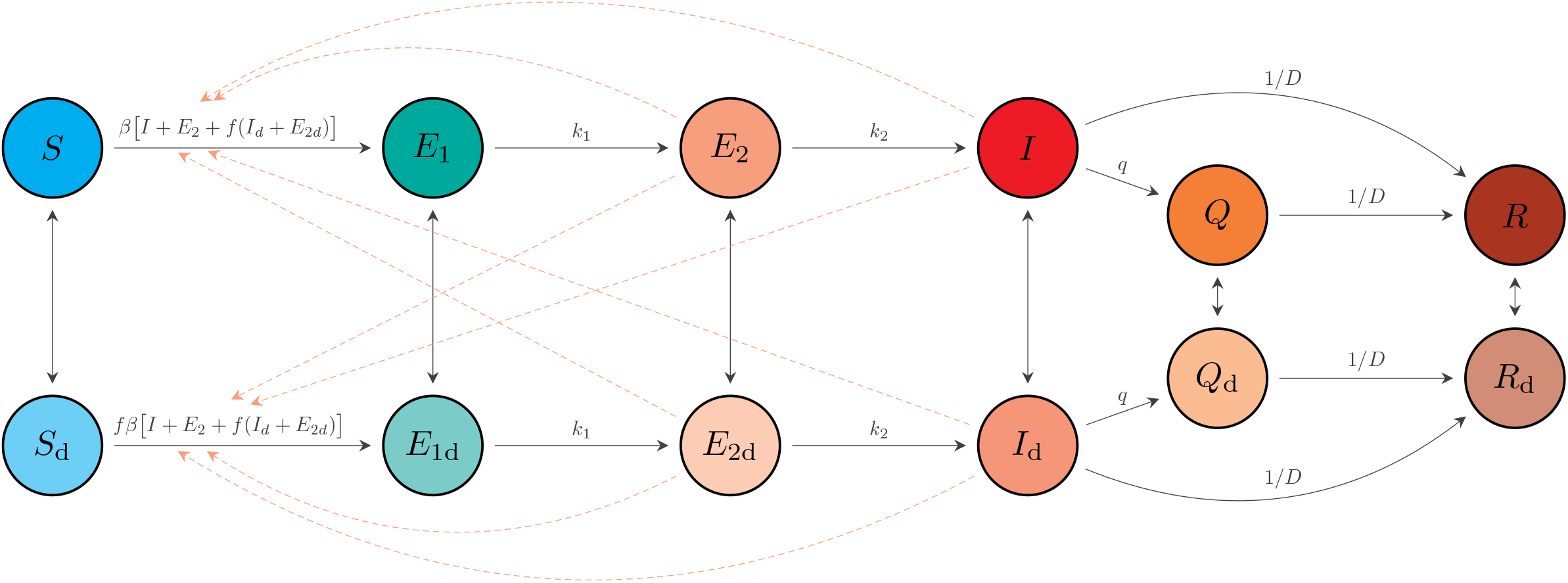
Schematic of the epidemiological model. Compartments: susceptible to the virus (*S*); exposed (*E*_1_); exposed, pre-symptomatic, and infectious (*E*_2_); symptomatic and infectious (*I*); quarantined (*Q*); and recovered or deceased (*R*). Recovered individuals are assumed to be immune. The model includes analogous variables for individuals practicing physical distancing: *S*_d_, *E*_1d_, *E*_2d_, *I*_d_, *Q*_d_, and *R*_d_. Solid arrows represent flow of individuals between compartments at rates indicated by the mathematical terms. Dashed lines show which compartments contribute to new infections. An individual in some compartment *X* can begin distancing and move to the corresponding compartment *X*_d_ at rate *u*_d_. The reverse transition occurs at rate *u*_*r*_. The model quickly settles on a fraction *e* = *u*_*d*_*/*(*u*_*d*_ + *u*_*r*_) participating in distancing, and dynamics depend on this fraction, rather than on the rates *u*_*d*_ and *u*_*r*_. Duplicated from Ref. [19] for clarity.

The model describes the time dynamics of susceptible (*S*), exposed (*E*_1_) exposed and infectious (*E*_2_), symptomatic and infectious (*I*), quarantined (*Q*) and recovered or deceased (*R*) individuals (see Fig. 4). It assumes that recovered individuals are immune to the virus. The model has analogous states for individuals practicing physical distancing, given by *S*_d_, *E*_1d_, *E*_2d_, *I*_d_, *Q*_d_, and *R*_d_. Physical distancing is implemented by reducing the contact rate, thereby lowering the spread of the virus. The model is fitted separately for each jurisdiction.

The system of differential equations for the non-physical-distancing population is given by:

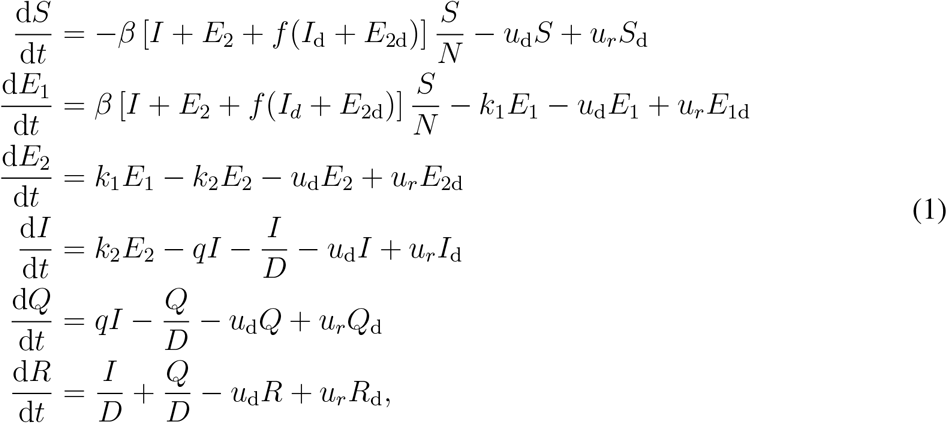

where *β* is the transmission rate, *f* is the physical distancing parameter, *D* is the average infectious period, *u*_d_ and *u*_*r*_ are the rates individuals move to and from the physical distancing compartments, *k*_1_ is the rate of moving from *E*_1_ to *E*_2_, *k*_2_ is the rate of moving from *E*_2_ to *I*, and *q* is the quarantine rate for movement from compartment *I* to *Q* [19]. In the model without interventions (neither distancing nor quarantine), the basic reproductive number *R*_0b_ is *β*(*D* + 1*/k*_2_), namely the transmission rate times the mean duration of the infectious state period. We explicitly estimate *R*_0b_ not *β*, and so *β* is given by *β* = *k*_2_*R*_0b_*/*(*Dk*_2_ + 1). The analogous system of equations for the physical-distancing population is given by

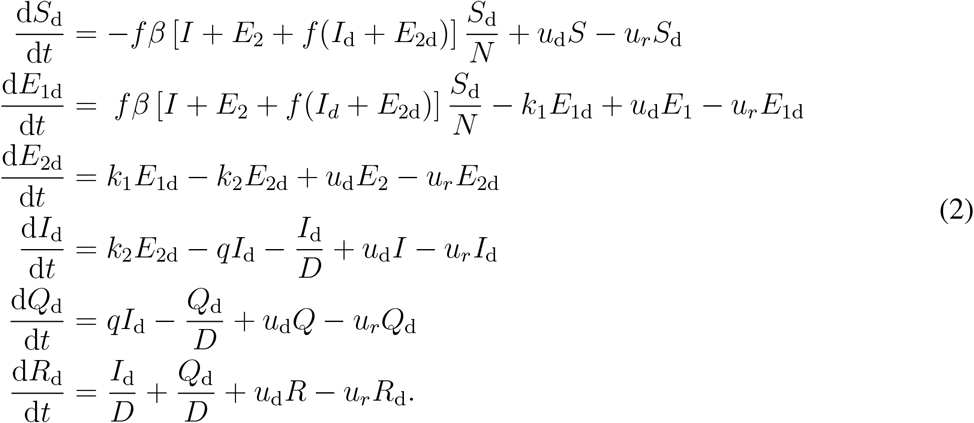

The force of infection for this population is a fraction *f* of that of the non-distancing population Eq. (1). In addition, note that the factor *f* appears twice in the force of infection. This is due to the fact that physical distancing helps in reducing the rate that “distancers” move about and contact others, and the rate at which they are contacted by anyone (distancing or otherwise) who is experiencing population contact. This factor changes with time to model the introduction and strength of NPI measures that reduce contact rates:

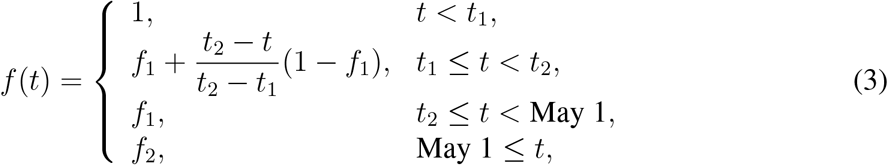

where *t*_1_ and *t*_2_ are the start and end times of the initial implementation of physical distancing measures such that *f* declines from 1 to *f*_1_ during this period, and *f*_2_ is the value of *f* after May 1 as physical distancing starts potentially relaxing. For each jurisdiction, *t*_1_, *t*_2_, *f*_1_, and *f*_2_ are estimated (see below).

Our overall approach is to estimate *f*_1_ and *f*_2_ using Bayesian inference. We also estimate the fraction of the population *e* = *u*_*r*_*/*(*u*_*d*_ + *u*_*r*_) engaged in NPI or distancing, the times *t*_1_ and *t*_2_, the and starting introduction size (prevalence at the model starting time). We use data from reported cases, despite the issues inherent in this [18], and compensate for variable testing through time where possible (see below) and for the delay between symptom onset and case reporting.

### Reported cases and testing model

We let *C*_*r*_ denote the number of recorded cases on day *r*. The number of people who become symptomatic on a given day *n* is the number moving from the exposed pre-symptomatic (*E*_2_ and *E*_2d_) to the symptomatic (*I* and *I*_d_) compartments, namely 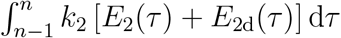. The expected number of reported cases on day *r* is a weighted sum of those who become symptomatic in previous days, where the weights are determined by the the delay between symptom onset and reporting [19]:

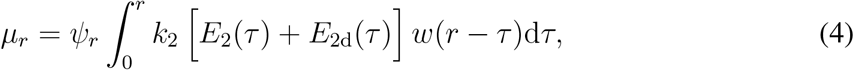

where *ψ*_*r*_ represents the sampling fraction on day *r* and we use a Weibull distribution with shape *k*_MLE_ and scale *λ*_MLE_ for *w*(·). If *ψ*_*r*_ = 1, then all estimated infectious people are tested and then become reported cases; *ψ*_*r*_ < 1 represents a reduction in expected cases on day *r* due to not everyone being tested. See Ref. [19] for further details on fitting *w*(·) from data. We used *k*_MLE_ and *λ*_MLE_ as estimated for British Columbia in Ref. [19] for the other regions (due to a lack of the necessary data), except for New Zealand for which A. Lustig and M. Plank (pers. comm.) fitted non-public data using our code [20].

Model fitting was performed in Stan with the R package ‘covidseir’ [21]. Code to reproduce the analysis is available at https://github.com/carolinecolijn/leeway-reopen-covid19.

## Data Availability

All data are publicly available. Code is provided on github.

## Acknowledgements

This work was supported by funding from the Michael Smith Foundation for Health Research and from Genome BC (project code COV-142). C.C. and J.S. are funded by the Federal Government of Canada’s Canada 150 Research Chair program. We thank Fisheries and Oceans Canada for their support.

## Supplementary Information

### Data processing and regional fitting

We obtained reported case data from publicly available sources (Table S1). We used baseline values of constant testing (0.2) where we were not aware of testing data indicating widened testing eligibility or steep increases in testing volume. Where these were indicated we increased the sample fraction accordingly (see Table S2). A number of jurisdictions also required additional data processing and/or modification to obtain regional fits:

- Several jurisdictions showed a strong weekly pattern in case reporting. In these cases (Belgium, Germany, Japan, Washington), we implemented a 3-day running average.
- Quebec public health officials announced that a computer error resulted in 1,317 missing positive COVID-19 cases between April 2nd–30th. As a result, we removed these 1,317 cases from the day they were eventually reported, and redistributed them evenly across days April 2nd–30th
- April 1st was an outlier in the number of observed cases in Ontario, seeing more than double the number of cases than other days during that week. To account for this, we redistributed the difference in the number of cases between April 1st and April 2nd evenly across the 5 days prior to April 1st.
- In the UK, April 11th was an outlier in the daily number of observed cases with almost double any other day during the pandemic so far. We redistributed the difference in the number of cases between April 10th and April 11th evenly across the 5 days prior to April 11th. The UK also made a large increase in the daily number of completed tests from April 30th onward, in line with the introduction of a government target of 100,000 tests per day by the end of April. We account for this in the model by increasing the assumed sampling fraction from 0.2 to 0.3 for April 30th onward.
- For British Columbia, we defined two change-points for the fraction of cases sampled. From March 1st–14th the sampling fraction was set to 0.14, from March 14th to April 21st it was set to 0.21 and from April 21st onward it was set to 0.37. These were obtained from a separate model fit that included daily hospitalizations and an assumed hospitalization fraction of 0.08.
- For New Zealand, we used a fixed sampling fraction of 0.4 under the assumption of a higher level of case detection. We also removed all cases arising from international travel.

**Table S1:** Publicly available data sources for reported COVID-19 cases across jurisdictions. See Fig. 2 for jurisdiction abbreviations.

| Jurisdiction | Data source URL |
| --- | --- |
| BC | http://www.bccdc.ca/health-info/diseases-conditions/covid-19/data |
| BE | https://epistat.wiv-isp.be/Covid/ |
| CA | https://covidtracking.com/ |
| DE | https://opendata.ecdc.europa.eu/covid19 |
| JP | https://ourworldindata.org/coronavirus |
| NY | https://covidtracking.com/api/v1/states/daily.csv |
| NZ | https://www.health.govt.nz/our-work/diseases-and-conditions/covid-19-novel-coronavirus |
| ON | https://github.com/ishaberry/Covid19Canada/ |
| QC | https://github.com/ishaberry/Covid19Canada/ |
| SE | https://opendata.ecdc.europa.eu/covid19 |
| UK | https://github.com/tomwhite/covid-19-uk-data |
| WA | https://covidtracking.com/ |

**Table S2:** Regional modelling initialization, data properties, and priors. Population numbers were obtained from local government websites. We set sampling fractions to 0.2 in most cases except for in British Columbia (BC) where the sampling fractions represent means from a fitted model that also accounts for daily hospitalizations with an assumed hospitalization fraction of 0.08 (changes on March 14, April 11, and April 21 due to known policy changes); New Zealand (NZ) where we assumed a higher sampling fraction; and the United Kingdom (UK) where there was a large increase in the daily number of completed tests from April 30 onward. The assumed sampling fraction should only affect modelled prevalence until substantial immunity is built up. See Fig. 2 for jurisdiction abbreviations.

| Detail | BC | BE | CA | DE | JP | NY | NZ | ON | QC | SE | UK | WA |
| --- | --- | --- | --- | --- | --- | --- | --- | --- | --- | --- | --- | --- |
| Data start | Mar 1 | Mar 3 | Mar 5 | Mar 1 | Mar 1 | Mar 5 | Mar 15 | Mar 1 | Mar 1 | Mar 1 | Mar 1 | Mar 1 |
| Data end | Jun 4 | Jun 6 | Jun 7 | Jun 7 | Jun 7 | Jun 7 | May 26 | Jun 7 | Jun 7 | Jun 8 | Jun 8 | Jun 3 |
| Prior mean for $t_1$ | Mar 16 | Mar 11 | Mar 8 | Mar 13 | Mar 27 | Mar 9 | Mar 18 | Mar 9 | Mar 10 | Mar 7 | Mar 12 | Mar 9 |
| Prior mean for $t_2$ | Mar 23 | Mar 21 | Mar 25 | Mar 22 | May 4 | Mar 28 | Mar 26 | Mar 24 | Mar 25 | Mar 28 | Mar 28 | Mar 29 |
| Prior sd for $t_1$ and $t_2$ | 0.1 | 0.1 | 0.1 | 0.1 | 0.1 | 0.1 | 0.2 | 0.1 | 0.1 | 0.2 | 0.1 | 0.1 |
| Prior mean for $e$ | 0.8 | 0.8 | 0.8 | 0.8 | 0.8 | 0.8 | 0.9 | 0.8 | 0.8 | 0.8 | 0.8 | 0.8 |
| Delay shape | 1.73 | 1.73 | 1.73 | 1.73 | 1.73 | 1.73 | 1.53 | 1.73 | 1.73 | 1.73 | 1.73 | 1.73 |
| Delay scale | 9.85 | 9.85 | 9.85 | 9.85 | 9.85 | 9.85 | 7.83 | 9.85 | 9.85 | 9.85 | 9.85 | 9.85 |
| Sampling fraction(s) | 0.14,<br>0.21,<br>0.37 | 0.2 | 0.2 | 0.2 | 0.2 | 0.2 | 0.4 | 0.2 | 0.2 | 0.2 | 0.2, 0.3 | 0.2 |
| Log mean for $I_0$ prior | 2.08 | 0 | 0 | 1.61 | 2.64 | 0 | -4.61 | 0 | 0 | 0 | 0 | 0 |
| $N$: population (millions) | 5.10 | 11.48 | 39.51 | 83.00 | 126.00 | 19.45 | 4.95 | 14.50 | 14.50 | 10.34 | 66.40 | 7.60 |

### Mobility data

We informed the priors for the start and end dates for physical distancing measures using Google mobility data [22] (Fig. S1). For each region, we use the daily average of the available public transportation data. We then fit a piecewise linear regression with two breakpoints to the data from each location using the R package ‘segmented’ [23]. We use the two fitted breakpoints (rounded to the nearest day) as prior means on the start and end dates *t*_1_ and *t*_2_ for each jurisdiction (Table S2). Two exceptions to this were New Zealand and Sweden. In New Zealand, mobility data suggested start and end dates of March 18th and March 30th, but an end date of March 26th was found to provide an improved fit. Similarly, in Sweden March 9th to March 20th were suggested by mobility data but this resulted in poor fit. Instead, we used March 6th to March 27th.

### Bayesian estimation

The joint posterior distribution given the case counts {*C*_*r*_} is

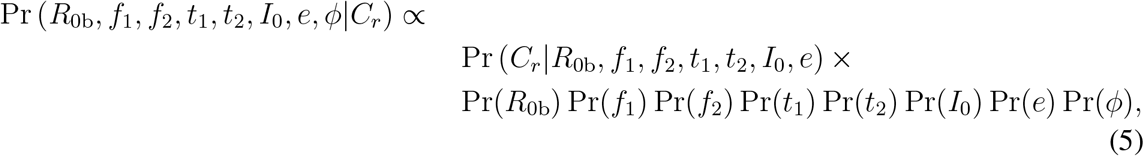

where *R*_0b_, *f*_1_, *f*_2_, *t*_1_, *t*_2_, *I*_0_, *e*, and *ϕ* are estimated parameters. We use a negative binomial likelihood for the observation component parameterized such that the variance scales quadratically with the mean [24]. We describe the parameters here for clarity: *R*_0b_ represents the basic reproductive number (without distancing but with quarantine), *f*_1_ and *f*_2_ represent the force of infection for the post-measures and recent periods, *I*_0_ represents the incidence at 30 days before the first day of data, *e* represents the fraction distancing, and *t*_1_ and *t*_2_ represent the dates that physical distancing starts and finishes ramping in, *ϕ* represents the (inverse) dispersion parameter.

We fit our models with Stan 2.19.3 [25,26] and R 3.6.2 [27] using our R package ‘covidseir’ [21]. We sampled from 4 chains with 400 iterations per chain and discarded the first half of each chain as warm-up. We initialized the chains at random values drawn from the priors. We assessed chain convergence with trace plots and via 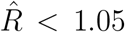 (the potential scale reduction factor) and ESS > 200 (the effective sample size) [26].

**Figure S1:**
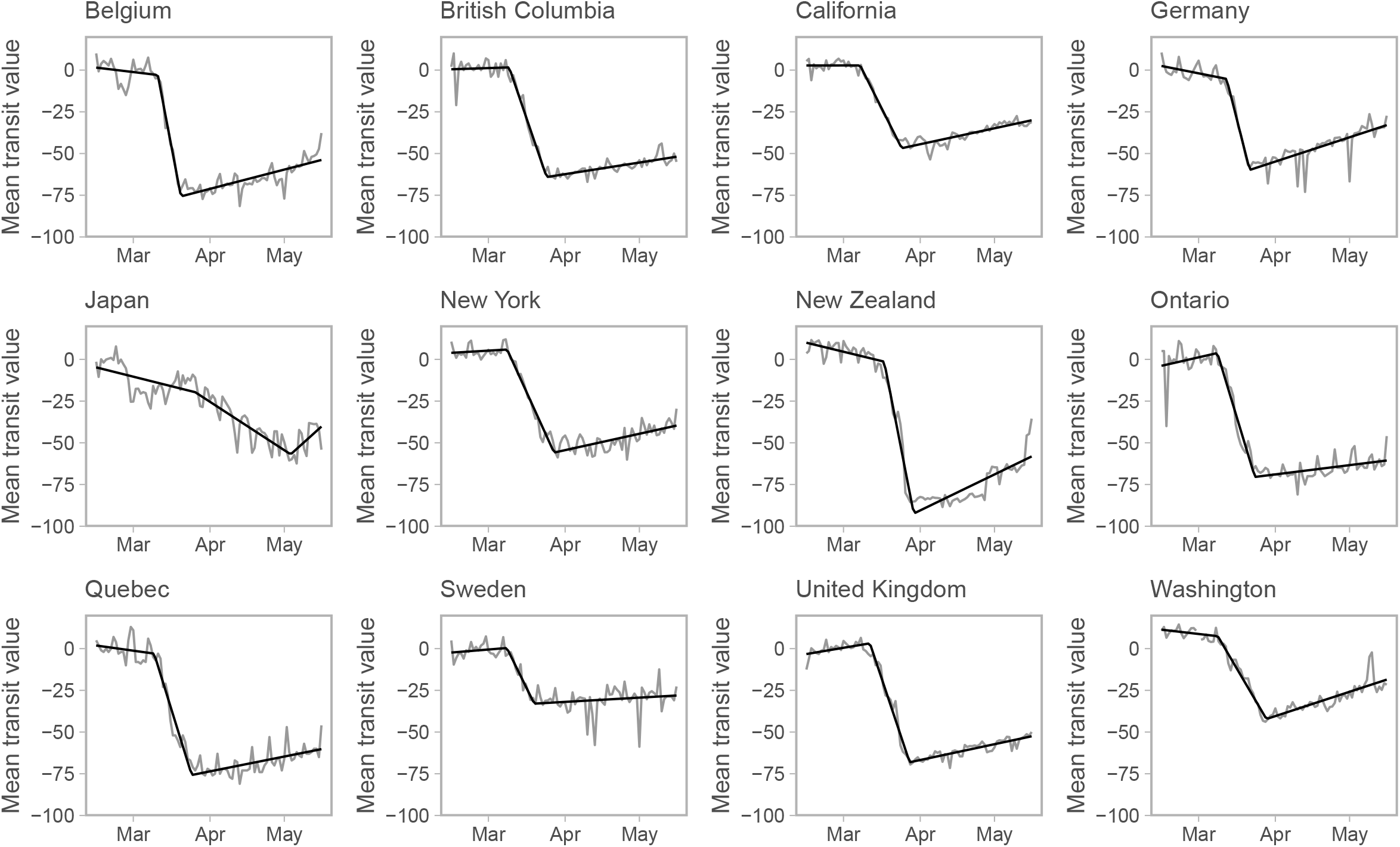
Mobility data from Ref. [22]. We use daily averaged public transit station percent change from baseline data for each jurisdiction. We fit a piecewise linear regression with two breakpoints using the R package ‘segmented’ [23] to inform the prior distributions for the timing of physical distancing.

**Table S3:** Fixed parameter values that are the same for all jurisdictions.

| Symbol | Definition | Default value | Justification |
| --- | --- | --- | --- |
| $D$ | Mean duration of the infectious period | 5 days | [28, 29] |
| $k_1$ | (time to infectiousness)$^{-1}$ ($E_1$ to $E_2$) | 0.2 days$^{-1}$ | [30–32] |
| $k_2$ | (time period of pre-symptomatic transmissibility)$^{-1}$ ($E_2$ to $I$) | 1 days$^{-1}$ | [31, 32] |
| $q$ | Quarantine rate | 0.05 | [33] |
| $u_r$ | Rate of people returning from physical distancing | 0.02 | [34] |

**Table S4:** Prior distributions for all jurisdictions; note that *I*_0_ and *e* have jurisdiction-dependent means (Table S2).

| Symbol | Definition | Prior distribution | Justification |
| --- | --- | --- | --- |
| $I_0$ | Number of infected people at an initial point in time | Lognormal with sd 1 | Small early introductions |
| $e$ | Proportion practicing distancing | Beta with sd 0.05 | Widespread measures |
| $R_{0b}$ | Basic reproductive number without distancing | Lognormal(log 2.6, 0.2) | [35, 36] |
| $f_1, f_2$ | Value of $f$ at different times (Eq. 3) | Beta with mean 0.4 and sd 0.2 | Weakly informative prior |
| $\phi$ | Inverse dispersion parameter of negative binomial observation model | $1/\sqrt{\phi} \sim \text{Normal}(0, 1)$ | [37] |

Some parameter values (Table S3) and prior distributions (Table S4) are the same for all jurisdictions, whereas some differ between jurisdictions (Table S2). The rate of people moving to physical distancing, *u*_d_, is calculated from the estimates of *e* = *u*_*r*_*/*(*u*_*d*_ + *u*_*r*_), because *e* is estimated and *u*_r_ is fixed. We used informative prior distributions on estimated parameters as follows (Table S4). The prior on *R*_0b_ encompasses values commonly published in the literature for SARS-CoV-2 [35, 36]. The prior for *f*_1_ and *f*_2_ results in a mean of 0.4 and a standard deviation of 0.2 to represent a moderately strong reduction in contact fraction while still being broad enough to encompass a wide range of values. We use lognormal priors for *t*_1_ and *t*_2_, with the means based on the piecewise regression analyses of the Google mobility data, and standard deviations of 0.1, except for New Zealand and Sweden for which less tight priors (standard deviation of 0.2) are needed. The prior on *ϕ* constrains the model to avoid substantial prior mass on a large amount of over-dispersion (small values of *ϕ*). The initial conditions of the state variables are defined consistently across jurisdictions, but depend on the each jurisdiction’s population size *N* and estimated values of *e* and *I*_0_ (Table S5). For most jurisdictions, the prior on *I*_0_ was set such that *I*_0_ has mean 1, representing a prior belief that the initial time point was set to a time without substantial numbers of cases in that location. New Zealand, where the total number of cases has been very small, required a smaller mean to obtain a satisfactory fit (0.01). For British Columbia we instead set the mean for the *I*_0_ prior to 8, as in [19], and for Germany and Japan (both having a more substantial number of cases prior to the initial time point) we used a mean equal to the reported number of observed cases 30 days before the initial time point: 5 and 14, respectively. These are summarized in Table S2.

**Table S5:** Initial values of variables. The parameters *e* and *I*_0_ are estimated in the model separately for each jurisdiction.

| Non-distancing |  | Distancing |  |
| --- | --- | --- | --- |
| Variable | Initial value | Variable | Initial value |
| $S$ | $(1 - e)(N - I_0)$ | $S_d$ | $e(N - I_0)$ |
| $E_1$ | $0.4(1 - e)I_0$ | $E_{1d}$ | $0.4eI_0$ |
| $E_2$ | $0.1(1 - e)I_0$ | $E_{2d}$ | $0.1eI_0$ |
| $I$ | $0.5(1 - e)I_0$ | $I_d$ | $0.5eI_0$ |
| $Q$ | 0 | $Q_d$ | 0 |
| $R$ | 0 | $R_d$ | 0 |

To calculate the posterior distribution of the ratio between the contact rate and the threshold (i.e., Fig. 1A), we apply the projection and regression approach described in Ref. [19]. We use a projection period of 25 days and evaluate *f* values ranging from 0.3 to 0.8. We then determine the threshold value and compare it to the estimated *f*_1_ or *f*_2_ for every posterior sample independently.

**Figure S2:**
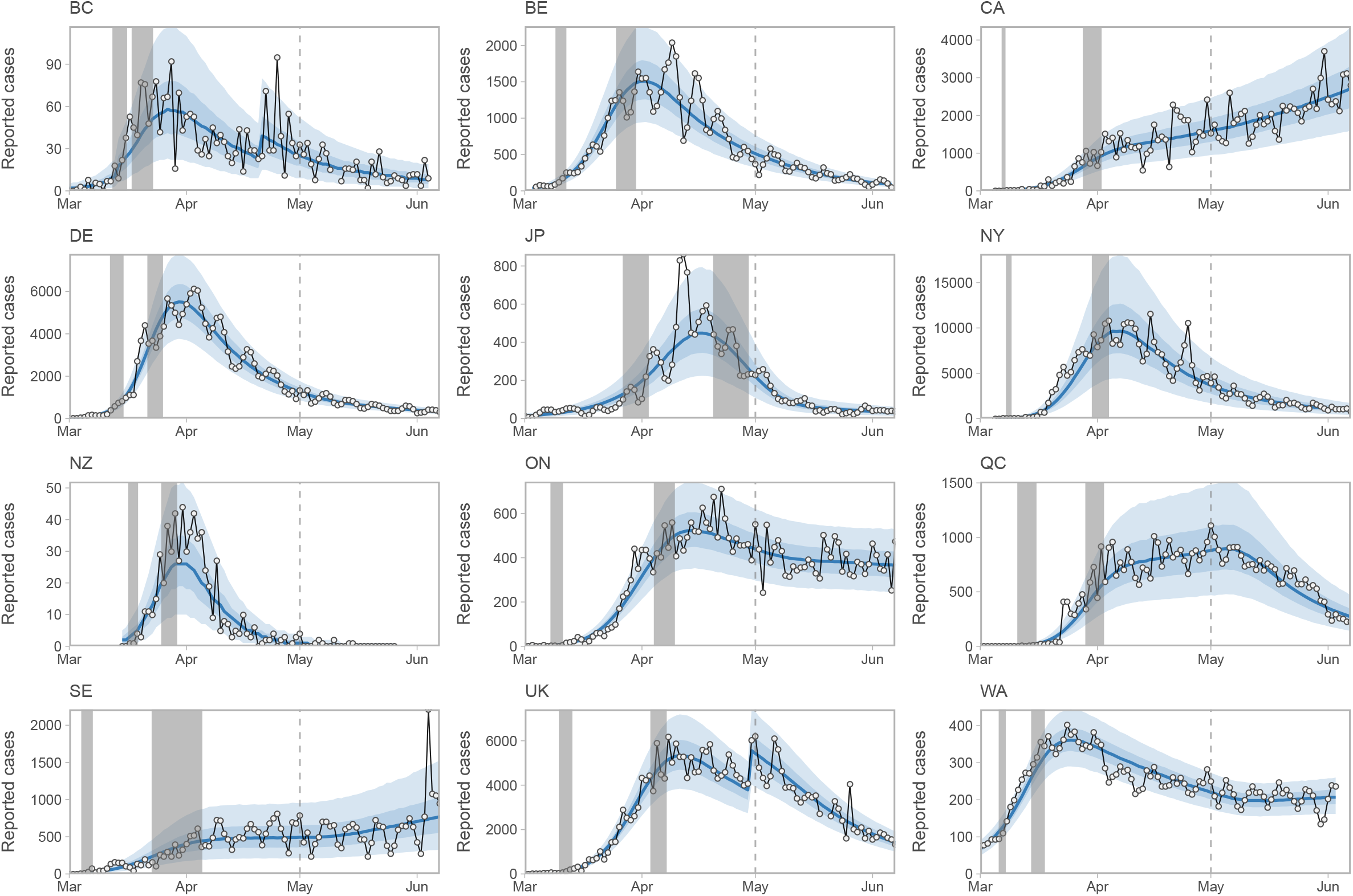
Reported case times series and model fits. These are the same as Fig. 1 but focused on the historical data model fits.

**Figure S3:**
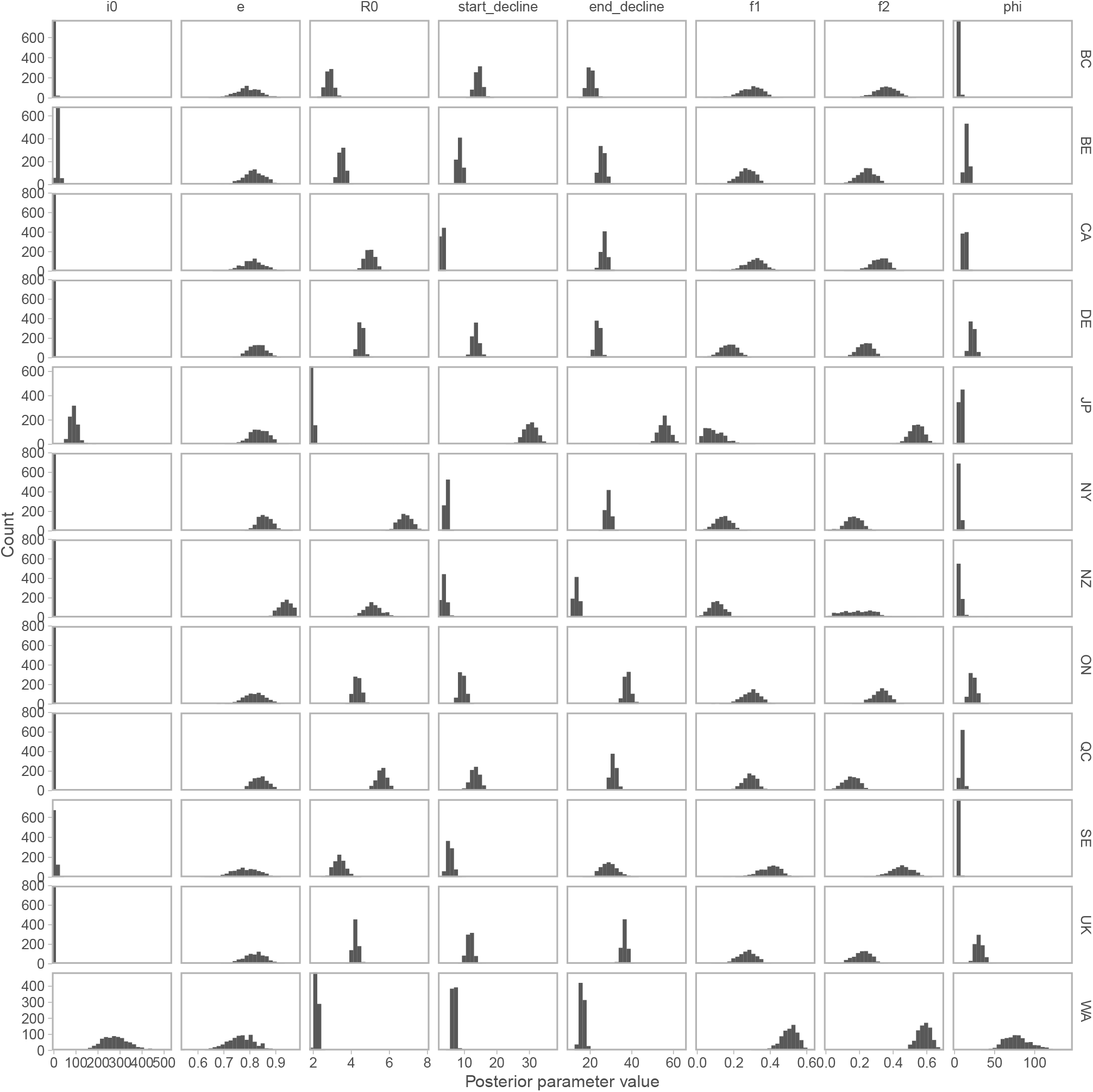
Posteriors for each jurisdiction of all estimated parameters from the Bayesian SEIR model. The columns “start decline” and “end decline” represent *t*_1_ and *t*_2_. *R*_0_ accounts for quarantine: *R*_0_ = *R*_0b_(1*/*(*q* + 1*/D*) + 1*/k*_2_)*/*(*D* + 1*/k*_2_)

**Figure S4:**
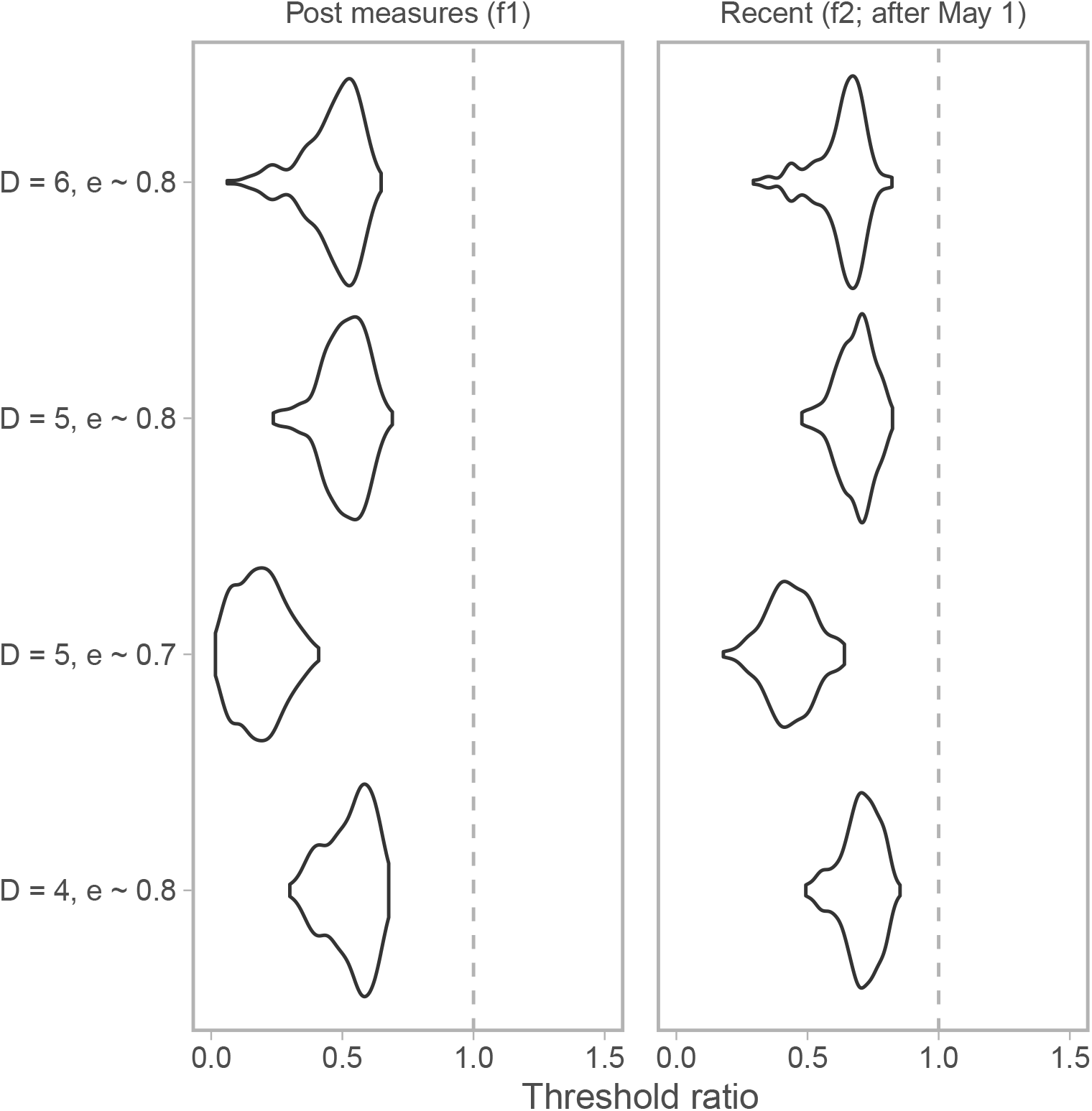
Example sensitivity for contact ratio in Germany to *D* (duration) of 4, 5, or 6 and *e* (fraction distancing) prior of mean 0.7 (and SD 0.025) or 0.8 (and SD of 0.05; as in the main models) for Germany.

